# Knowledge mobilization with and for equity-deserving communities invested in research: A scoping review protocol

**DOI:** 10.1101/2024.09.06.24313221

**Authors:** Ramy Barhouche, Samson Tse, Fiona Inglis, Debbie Chaves, Erin Allison, Tina Colaco, Melody E. Morton Ninomiya

## Abstract

The practice of putting research into action is known by various names, depending on disciplinary norms. Knowledge mobilization, translation, and transfer (collectively referred to as K*) are three common terminologies used in research literature. Knowledge-to-action opportunities and gaps in academic research often remain obscure to non-academic researchers in communities, policy and decision makers, and practitioners who could benefit from up-to-date information on health and wellbeing. Academic research training, funding, and performance metrics rarely prioritize or address non-academic community needs from research. We propose to conduct a scoping review on reported K* in community-driven research contexts, examining the governance, processes, methods, and benefits of K*, and mapping who, what, where, and when K* terminology is used. This protocol paper outlines our approach to gathering, screening, analyzing, and reporting on available published literature from four databases.

## Introduction

Bridging the gap between produced research knowledge and action, policy, and/or practice has been a common goal across disciplines for the past few decades, often involving synthesis, analysis, dissemination, and application (1). However, there has been some confusion in the use of various terms and definitions to represent this goal and its application process. In academia, for example, terms such as *knowledge translation*, *knowledge mobilization*, and *knowledge transfer* are widely utilized (2). Similarly, Canada’s *Tri-Agencies*, the three main federal funding agencies for research, employ varied terminology and definitions for disseminating research knowledge. For instance, the Social Sciences and Humanities Research Council (SSHRC) (3) defines knowledge mobilization as an umbrella term encompassing activities related to the production and use of research results, including synthesis, dissemination, transfer, exchange, and co-creation by researchers and knowledge users. The Canadian Institutes of Health Research (CIHR), on the other hand, use terms like knowledge translation, integrated knowledge translation (IKT), and end-of-grant knowledge translation, emphasizing a dynamic and iterative process of knowledge synthesis, dissemination, exchange, and ethically-sound application (4). The manuscript will adopt the term K* to collectively refer to these terms (5).

There has been a recent trend for utilizing research findings to inform practices, policies, and decisions through K* efforts. However, there is still a significant gap in research application, and knowledge derived from research has not effectively translated into health practices and policies (6). The gap between research findings and application can be attributed to sociocultural, organizational, and economic factors that play a significant role influencing the research and mobilization process (1,7). Furthermore, academics may lack readiness to engage in K* work, or if they do, their existing heavy workload limits their capacity to do so (8). This disconnect significantly impacts community members, who are often knowledge users.

Similarly, community-based participatory research (CBPR) is recognized as a shift in the research paradigm that can enhance the utilization of research knowledge by involving knowledge users more extensively in the research process (9). Like K*, CBPR also includes different aspects of shared and active community engagement in the research process, as well as enhancing community health through the integration of research and action (10). In particular, CBPR complements IKT by emphasizing a democratic process of co-creating knowledge that aligns closely with the needs of knowledge users (9). Thus, CBPR is essential for research that aims to enhance the health and wellbeing of individuals who are systematically marginalized or discriminated against. Traditional research has often been characterized by an unequal power dynamic between researchers and participants, a gap that CBPR can address (11). For example, Christensen (12) and Morton Ninomiya et al. (13) have demonstrated the effectiveness of CBPR in engaging Indigenous communities in a respectful and culturally responsive way that led to impactful research outcomes. The essence of K* lies in the significance it brings to research. To promote equity and sovereignty-deserving communities, it can be argued that it is ethically imperative to ensure that community-driven K* needs are integrated into CBPR.

### Objectives

With the diverse terminology and definitions used to describe the “knowledge-to-action” gap, there is understandable confusion and potential frustration regarding the operationalization and use of K* terms. A preliminary search of databases revealed no current or ongoing scoping reviews on this topic. The involvement of community members from equity seeking, deserving, or denied groups is essential for research committed to1) addressing health and wellness inequities faced by different communities and/or 2) “nothing about us without us”. We propose to conduct a scoping review study.

### Research Questions

The *scoping review* will answer the following questions about research aimed at supporting and serving non-academic equity and/or sovereignty-seeking/deserving/denied communities:

1. How are terms and concepts around knowledge mobilization (K*) being used, defined, and cited in published literature?
2. In what ways are community partners involved in K* priorities, planning, and efforts?
3. What are reported wellbeing-related impacts and outcomes being addressed within communities through K* efforts?

## Methods

### Search Strategy

This scoping review protocol was developed using the JBI Manual for Evidence Synthesis (14) and adheres to the PRISMA-ScR as developed by Tricco et al. (15). Our protocl is registered with Open Science Framework (OSF). As a team, we identified 11 papers (16–26) that met all inclusion criteria to use as a test set while we developed our search strategy. After extensive testing, refining, and discussions as a team about concepts and terminology, the initial Medline (ProQuest) search strategy was finalized by two academic librarians [DC, FI]. This search was then translated by [RB] for PsycINFO (ProQuest), CINAHL with Full Text (EBSCOhost), and Web of Science Core Collection (Clarivate).

The search strategy focuses on four primary concepts: vulnerable populations (i.e., equity and/or sovereignty-seeking/deserving/denied communities), community-based research, knowledge translation, and health and wellbeing. The search terms around the concept of “vulnerable populations” were adapted from four published search strategies (27–29). The terms around the concept of K* search strategy was adapted from Morton Ninomiya et al. (30). We limited the search by date, including publications in 2010 onwards due to the increased international discourse and interest in K* in health and wellness research beginning at that time. Searching for research with equity and/or sovereignty-seeking/deserving/denied communities, often framed as “vulnerable populations”, is challenging because it sometimes includes offensive and outdated terms that are exclusionary. These terms were included in the search strategy to ensure a comprehensive search. The authors acknowledge the harmful nature of these terms and inform the reader of their inclusion. We will conduct a backwards and forwards search of the references in 1) review and discussion papers that do not meet the inclusion criteria, and 2) all included papers after full-text screening – in search of additional K* terms that we may have missed. See supplementary file 1 for the full Medline search strategy.

### Inclusion and Exclusion Criteria

The scoping review will only include primary studies published in English in which authors identify 1) their understanding of K* terminology with a definition, description, or citation; 2) *how* academic researchers engaged with non-academic communities; 3) *what* K* efforts took place; and 4) *how* K* efforts impacted individual, family, and/or community wellbeing. We will exclude grey literature sources including reports, books, commentaries, and theses/dissertations from the scoping review. Because of the rise of international discourse and explicit interest in K* in health and wellness research that started around the 2010s, the date limit will be from 2010 to present. Only papers that are about equity and/or sovereignty seeking/deserving/denied communities will be included. The inclusion and exclusion criteria and their rationale are listed in Table 1 below.

**Table 1.** Inclusion/exclusion Criteria and Rationale.

| Title and Abstract Screening Phase |  |  |
| --- | --- | --- |
| Include | Exclude | Rationale |
| Printed in English. | Printed in languages other than English. | It is the only shared fluent language of the research team. |
| Published in 2010 or later. | Published before 2010 or older. | We want the most updated, especially considering the rapid changes in KT literature. We are interested in seeing how it has been developed in the past 15 years or so. Google trends show consistent usage in search terms from 2010 and beyond. |
| Primary research papers, including case studies. | Discussion papers, autoethnographies, and reviews. | We want papers that are researching and applying K*. |
| Peer-reviewed and published. | Grey literature. | Interested in community-partnered academic research that often reflects disciplinary training, practice, language, and funding. For a global study, including grey literature would not be feasible. |
| Use specific terminology to describe K* in their paper. | Papers that do not use a specific term(s) to refer to K* initiatives, even if they describe elements of K*. | We are interested in terms people use to name and describe K* initiatives they facilitate. |
| The study addresses or supports something that aims to improve wellbeing for equity | Exclude studies focused on capitalistic endeavors for monetary gain and power, or focused on communities that | We are only interested in K* efforts on studies that aim to improve forms of wellbeing for communities |
| and/or sovereignty-deserving communities. | are not equity or sovereignty-deserving. | who do not have equitable access to health and wellbeing. |
| **Full-text screening (added criteria)** |  |  |
| Authors define or cite K* term(s) being used in the context of the study. | Papers that are unclear how the authors understand or use K* terminology. | This is an essential component of this study. |
| Authors state how non-academic community research partners were involved in the study, including K* related activities. Were involved in the study beyond being research participants. | Authors of the paper do not indicate how community was involved in the study, including K* related efforts. | Understanding how non-academic communities are involved in the research and K* are key to this study. |
| Authors state impacts and outcomes from K* efforts, in relation to non-academic research partner needs and priorities. | Papers that do not identify impacts and outcomes from K* efforts in relation to non-academic research partner needs and priorities. | The impacts and outcomes from K* effort on non-academic researchers is the focus of the scoping review. |

### Source of Evidence Selection

Search results will be imported into Covidence (31) and duplicate records will be removed automatically. To ensure high inter-rater reliability, four researchers [RB, JK, MM, ZP] involved in the screening process will screen the same 25 randomly selected records, based on the titles and abstracts. If we do not reach a minimum of 75% agreement, we will repeat the process with five more randomly selected records, until 75% agreement has been established.

Each title and abstract will be independently screened by two researchers. If one researcher thinks the record meets the inclusion criteria and the other does not, papers will be put in a “conflict” list. Each item in the conflict list will be resolved with a conversation between the two researchers and if consensus cannot be reached, the record will be included for full-text screening.

All papers included after the title and abstract screening will be uploaded for the full-text screening. Each full-text will be independently screened by two researchers and again, conflicts will be resolved through a conversation between the two researchers involved. If researchers cannot reach consensus, senior researcher [MMN] will make the final decision. Reasons for full-text exclusion will be tracked within Covidence and reported in the results. Only papers that met the inclusion criteria based on the full-text screening will be included for data extraction and analysis.

### Data Extraction and Analysis

#### Data Extraction

The scoping review extraction fields (Table 2) was developed by ST, RB, and MMN to record key K* information from the literature that will be selected, including author, reference, and relevant results aligned with the study’s research question (32). Data from all included full-texts will be manually extracted and recorded in Covidence (31). Two researchers will read through the K* literature and extract the data separately. The researchers will then meet to discuss and compare the extracted data to ensure accuracy and reliability.

**Table 2.** Data Extraction and Analysis.

| Category | Description |
| --- | --- |
| General Data | • Publication Year |
|  | • Location of study (region, country) |
|  | • Location of lead/senior researcher affiliation (region, country) |
|  | • Discipline (field of research and/or author affiliation) |
| Study | • Population studied/served/partnering |
|  | • Research approach, methodology & methods |
|  | • Research governance (e.g. role of community partner(s)) |
| K* | • Authorship inclusion of community involved<br>• K* terminology used<br>• K* definition(s) and/or citations<br>• Community partner involvement in K* prioritizing, planning, implementing<br>• Other non-researcher rights/stakeholders involved in K* prioritizing, planning, implementing<br>• Target group of K*<br>• K* aims<br>• Description of all K* efforts at planning & development stage<br>• Description of all K* efforts at implementing stage |
| Outcomes | • Reported impacts or outcomes<br>• Methods of tracking impacts and outcomes |

#### Data Analysis

We will use thematic analysis as described by Braun and Clarke (33,34) to identify, analyze, and report on themes within the open-ended extracted data. The analysis of extracted open-ended and descriptive K* information and outcomes will be analyzed over six phases: (1) becoming familiar with the data, (2) generating initial codes, (3) searching for themes, (4) reviewing the themes, (5) defining and naming themes, and (6) producing the final report.

Once the extracted data is approved, the research team will each familiarize themselves with it and conduct an initial analysis independently before discussing and reaching a consensus on themes. After the consensus is reached, the team will re-analyze and revise the data results one more time before sharing them with the rest of the research team. Finally, in the sixth phase, the analysis findings will be incorporated into the results section, supported by quotations from the literature.

## Discussion

To our knowledge, this scoping review analysis is the first of its kind. Since 2010, there have been related reviews conducted on *integrated knowledge translation* with health policies (35,36) a realist evaluation of *knowledge transfer* implementation by health researchers (37); *knowledge translation* and its supported definitions, theories, models, and frameworks (38); and knowledge translation in Indigenous health research (39), for example. Our scoping review analysis will gather a broader disciplinary scope of literature and terminology than other reviews to date. Our scoping review is unique in that it will focus on studies where non-academic community partners are presented as being clearly invested and engaged in K* efforts – in ways that address inequities and support sovereignty, improving the health and wellness of equity-deserving/seeking/denied communities.

There is growing interest globally within academic and research institutions, research funding bodies, and government priorities to support research with equity-seeking/deserving/denied communities. Evidence to suggest the growing interest are reflected in university strategic plans, Truth and Reconciliation Calls to Action, targeted funding calls, and government public health frameworks, for example. Despite the increased attention to “applied research” with emancipatory goals, community-based researchers have been writing about limitations, challenges, and the inequitable nature of research funding timelines, accountabilities of researchers and funders to communities – for many years. Given that most scientific research is written, and therefore framed and curated, by academic researchers, it will be interesting to see how authors use K* terminology with vagueness to imply research “value” versus describing in compelling detail how community made informed decisions around K* priorities and activities.

We assert that research training programs and curriculum in disciplines invested in human health and wellbeing must educate, mentor, and model K*. That said, there is a dearth of training and research in the area of K* with equity or sovereignty-seeking/deserving/denied communities. The asymmetry between potential and actual transformation through research can be addressed, in part, by making exemplary K* research more visible and calling on authors of future papers study and report on how K* was planned, prioritized, implemented, and evidenced.

### Limitations

A limitation of the scoping review is the lack of uniform terminology on the topic of K*, across disciplines, communities, and geographies. As the topic of K* for research involving equity and sovereignty-seeking/deserving/denied communities remains unexplored, there are no consistent subject headings for the research team to reference or use as a guide for the literature searches. Our analysis will be limited by the keywords we have identified as relevant and introduces bias, as we elucidate additional K* terms being used. We aim to minimize this bias by conducting a backwards and forwards search of the references in review and discussion papers – in search of additional K* terms that we may have missed. In addition, limiting the scoping review to English could create a limitation to explore non-English literature, since this paper is aiming for a global understanding of these terms.

Our study only includes peer reviewed published literature and therefore excludes grey literature such as study reports, theses and dissertations where K* may be defined, discussed, and described. We excluded grey literature to keep the scoping review feasible. Including grey literature would be unwieldy, given that our scoping review is global and inter/multidisciplinary. It would be near impossible to create a comprehensive and strategic grey literature search unless we severely limited the search by geography, discipline, or date however, we do not have a clear reason to do so. Considering that K* language carries the most currency in academic research and research funding cultures, we feel that limiting the search to peer-review literature is justified.

For the scoping review portion of our study, we will exclude papers that do not report on outcomes or impacts from K* efforts. Unless a study explicitly includes tracking, reporting, or observing outcomes and impacts from the K* efforts, authors may submit their manuscripts to journals before K* impacts and outcomes are known – and – the life cycle of most research grants are too short to include studying K* impacts and outcomes.

## Conclusion

The combination of using a scoping review to gather, examine, and visualize literature reporting on K* terminology, definitions, and priorities in research with equity or sovereignty-seeking/deserving/denied communities globally will paint an aerial view and nuanced summary of impactful research. Our study findings will be used to draw attention to examples of transformative research as well as reveal patterns of K* activities, outcomes, ethics, and accountabilities across geographies and disciplines.

## Data Availability

No datasets were generated or analysed during the current study. All relevant data from this study will be made available upon study completion.

## Acknowledgements

RB, ST,FI, DC, MEMN contributed to the conceptualization; RB and ST shared project administration; RB, ST, MEMN contributed to the original manuscript drafts; FI and DC facilitated the validation; all but AE and TC were involved in the review and editing of the manuscripts though all co-authors read the manuscript before submission; and all co-authors were involved in the methodology.

## Supplementary File 1 – The Search Strategy in Medline (ProQuest)

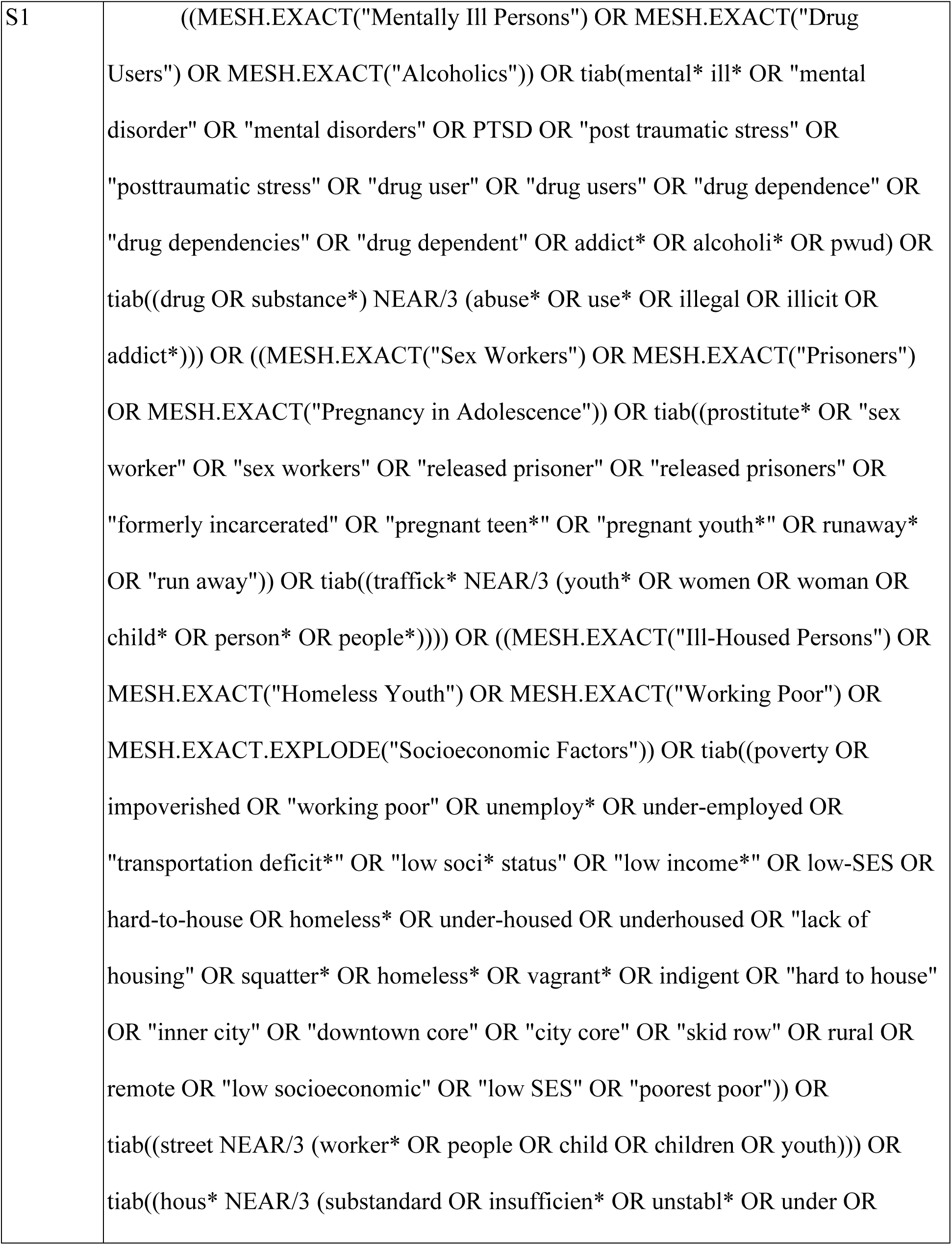
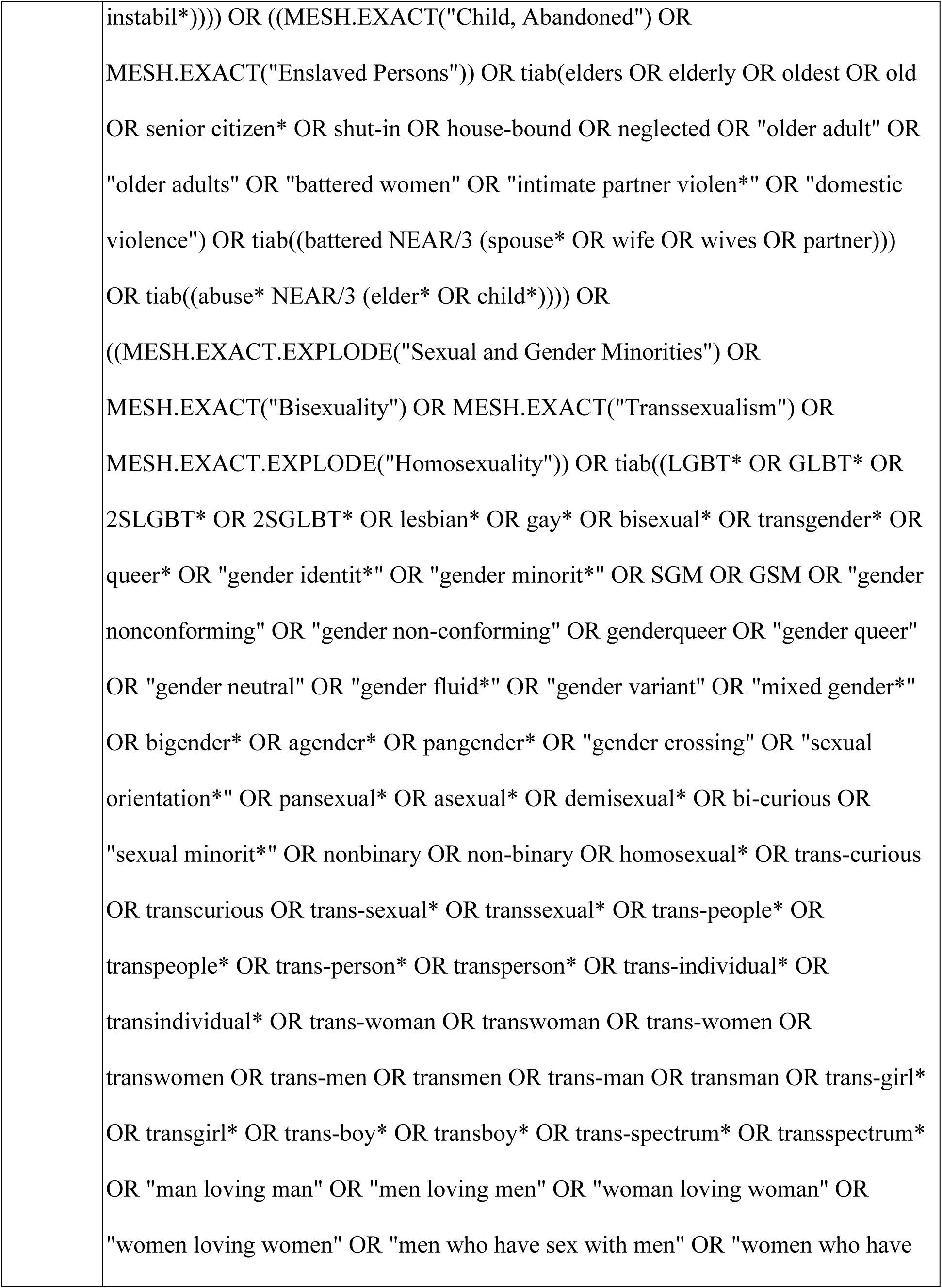
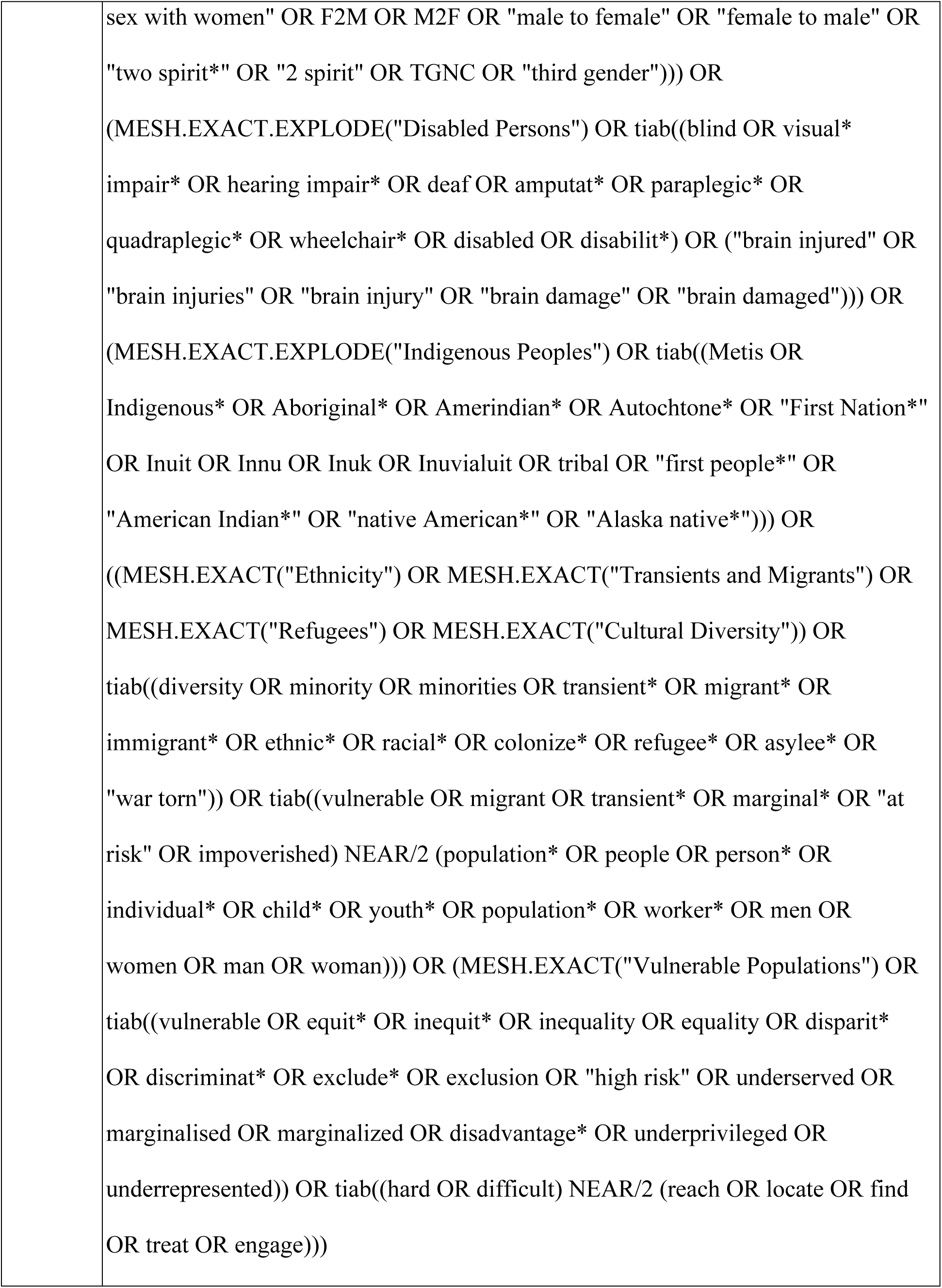
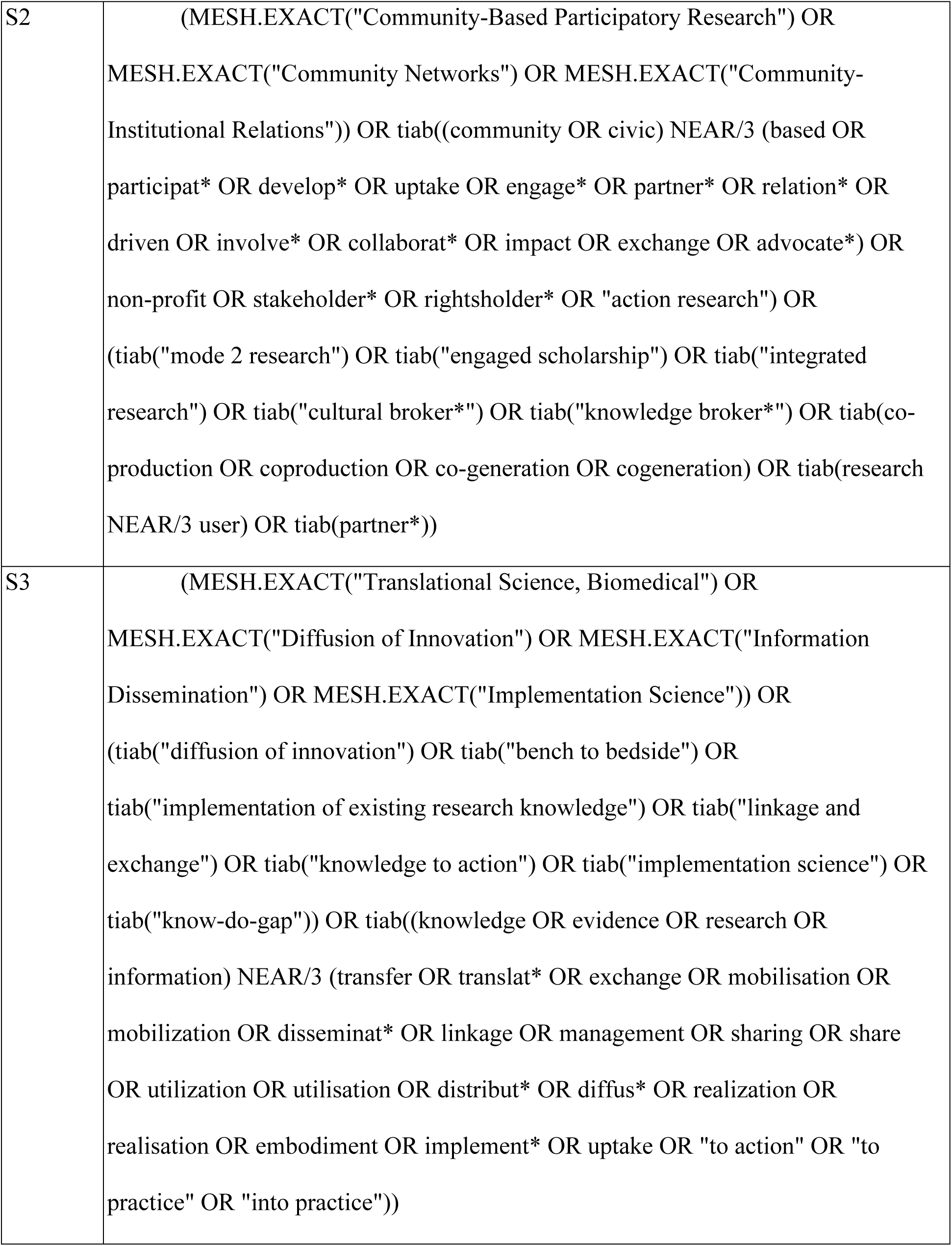
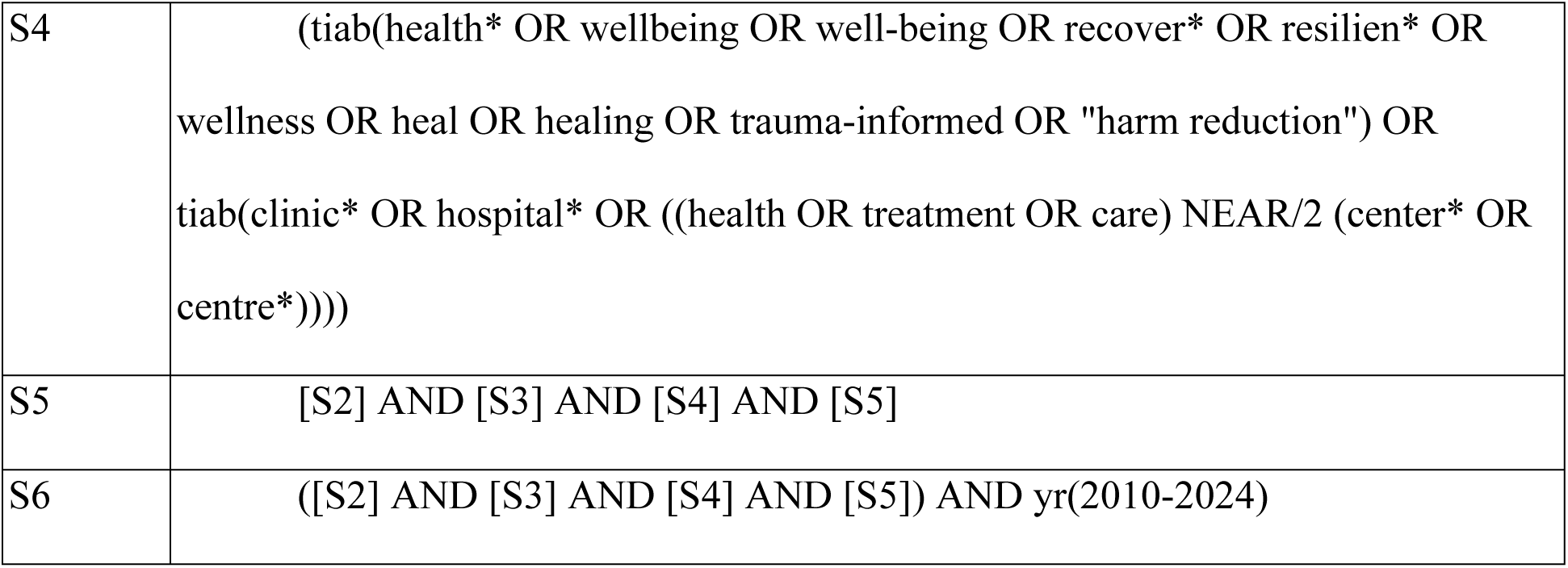

|  |  |
| --- | --- |
| S1 | ((MESH.EXACT("Mentally Ill Persons") OR MESH.EXACT("Drug Users") OR MESH.EXACT("Alcoholics")) OR tiab(mental* ill* OR "mental disorder" OR "mental disorders" OR PTSD OR "post traumatic stress" OR "posttraumatic stress" OR "drug user" OR "drug users" OR "drug dependence" OR "drug dependencies" OR "drug dependent" OR addict* OR alcoholi* OR pwud) OR tiab((drug OR substance*) NEAR/3 (abuse* OR use* OR illegal OR illicit OR addict*))) OR ((MESH.EXACT("Sex Workers") OR MESH.EXACT("Prisoners") OR MESH.EXACT("Pregnancy in Adolescence")) OR tiab((prostitute* OR "sex worker" OR "sex workers" OR "released prisoner" OR "released prisoners" OR "formerly incarcerated" OR "pregnant teen*" OR "pregnant youth*" OR runaway* OR "run away"))) OR tiab((traffick* NEAR/3 (youth* OR women OR woman OR child* OR person* OR people*))) OR ((MESH.EXACT("Ill-Housed Persons") OR MESH.EXACT("Homeless Youth") OR MESH.EXACT("Working Poor") OR MESH.EXACT.EXPLODE("Socioeconomic Factors")) OR tiab((poverty OR impoverished OR "working poor" OR unemploy* OR under-employed OR "transportation deficit*" OR "low soci* status" OR "low income*" OR low-SES OR hard-to-house OR homeless* OR under-housed OR underhoused OR "lack of housing" OR squatter* OR homeless* OR vagrant* OR indigent OR "hard to house" OR "inner city" OR "downtown core" OR "city core" OR "skid row" OR rural OR remote OR "low socioeconomic" OR "low SES" OR "poorest poor"))) OR tiab((street NEAR/3 (worker* OR people OR child OR children OR youth))) OR tiab((hous* NEAR/3 (substandard OR insufficien* OR unstabl* OR under OR |

|  |
| --- |
| instabil*)))) OR ((MESH.EXACT("Child, Abandoned") OR<br>MESH.EXACT("Enslaved Persons")) OR tiab(elders OR elderly OR oldest OR old<br>OR senior citizen* OR shut-in OR house-bound OR neglected OR "older adult" OR<br>"older adults" OR "battered women" OR "intimate partner violen*" OR "domestic<br>violence") OR tiab((battered NEAR/3 (spouse* OR wife OR wives OR partner)))<br>OR tiab((abuse* NEAR/3 (elder* OR child*)))) OR<br>((MESH.EXACT.EXPLODE("Sexual and Gender Minorities") OR<br>MESH.EXACT("Bisexuality") OR MESH.EXACT("Transsexualism") OR<br>MESH.EXACT.EXPLODE("Homosexuality")) OR tiab((LGBT* OR GLBT* OR<br>2SLGBT* OR 2SGLBT* OR lesbian* OR gay* OR bisexual* OR transgender* OR<br>queer* OR "gender identit*" OR "gender minorit*" OR SGM OR GSM OR "gender<br>nonconforming" OR "gender non-conforming" OR genderqueer OR "gender queer"<br>OR "gender neutral" OR "gender fluid*" OR "gender variant" OR "mixed gender*"<br>OR bigender* OR agender* OR pangender* OR "gender crossing" OR "sexual<br>orientation*" OR pansexual* OR asexual* OR demisexual* OR bi-curious OR<br>"sexual minorit*" OR nonbinary OR non-binary OR homosexual* OR trans-curious<br>OR transcurious OR trans-sexual* OR transsexual* OR trans-people* OR<br>transpeople* OR trans-person* OR transperson* OR trans-individual* OR<br>transindividual* OR trans-woman OR transwoman OR trans-women OR<br>transwomen OR trans-men OR transmen OR trans-man OR transman OR trans-girl*<br>OR transgirl* OR trans-boy* OR transboy* OR trans-spectrum* OR transspectrum*<br>OR "man loving man" OR "men loving men" OR "woman loving woman" OR<br>"women loving women" OR "men who have sex with men" OR "women who have |
| sex with women" OR F2M OR M2F OR "male to female" OR "female to male" OR "two spirit*" OR "2 spirit" OR TGNC OR "third gender")) OR<br>(MESH.EXACT.EXPLODE("Disabled Persons") OR tiab((blind OR visual* impair* OR hearing impair* OR deaf OR amputat* OR paraplegic* OR quadraplegic* OR wheelchair* OR disabled OR disabilit*) OR ("brain injured" OR "brain injuries" OR "brain injury" OR "brain damage" OR "brain damaged")))) OR<br>(MESH.EXACT.EXPLODE("Indigenous Peoples") OR tiab((Metis OR Indigenous* OR Aboriginal* OR Amerindian* OR Autochtone* OR "First Nation*" OR Inuit OR Innu OR Inuk OR Inuvialuit OR tribal OR "first people*" OR "American Indian*" OR "native American*" OR "Alaska native*")))) OR<br>((MESH.EXACT("Ethnicity") OR MESH.EXACT("Transients and Migrants") OR MESH.EXACT("Refugees") OR MESH.EXACT("Cultural Diversity")) OR tiab((diversity OR minority OR minorities OR transient* OR migrant* OR immigrant* OR ethnic* OR racial* OR colonize* OR refugee* OR asylee* OR "war torn")) OR tiab((vulnerable OR migrant OR transient* OR marginal* OR "at risk" OR impoverished) NEAR/2 (population* OR people OR person* OR individual* OR child* OR youth* OR population* OR worker* OR men OR women OR man OR woman))) OR (MESH.EXACT("Vulnerable Populations") OR tiab((vulnerable OR equit* OR inequit* OR inequality OR equality OR disparit* OR discriminat* OR exclude* OR exclusion OR "high risk" OR underserved OR marginalised OR marginalized OR disadvantage* OR underprivileged OR underrepresented)) OR tiab((hard OR difficult) NEAR/2 (reach OR locate OR find OR treat OR engage))) |
| S2 | (MESH.EXACT("Community-Based Participatory Research") OR MESH.EXACT("Community Networks") OR MESH.EXACT("Community-Institutional Relations")) OR tiab((community OR civic) NEAR/3 (based OR participat* OR develop* OR uptake OR engage* OR partner* OR relation* OR driven OR involve* OR collaborat* OR impact OR exchange OR advocate*) OR non-profit OR stakeholder* OR rightsholder* OR "action research") OR (tiab("mode 2 research") OR tiab("engaged scholarship") OR tiab("integrated research") OR tiab("cultural broker*") OR tiab("knowledge broker*") OR tiab(co-production OR coproduction OR co-generation OR cogeneration) OR tiab(research NEAR/3 user) OR tiab(partner*)) |
| S3 | (MESH.EXACT("Translational Science, Biomedical") OR MESH.EXACT("Diffusion of Innovation") OR MESH.EXACT("Information Dissemination") OR MESH.EXACT("Implementation Science")) OR (tiab("diffusion of innovation") OR tiab("bench to bedside") OR tiab("implementation of existing research knowledge") OR tiab("linkage and exchange") OR tiab("knowledge to action") OR tiab("implementation science") OR tiab("know-do-gap")) OR tiab((knowledge OR evidence OR research OR information) NEAR/3 (transfer OR translat* OR exchange OR mobilisation OR mobilization OR disseminat* OR linkage OR management OR sharing OR share OR utilization OR utilisation OR distribut* OR diffus* OR realization OR realisation OR embodiment OR implement* OR uptake OR "to action" OR "to practice" OR "into practice")) |
| S4 | (tiab(health* OR wellbeing OR well-being OR recover* OR resilien* OR wellness OR heal OR healing OR trauma-informed OR "harm reduction") OR tiab(clinic* OR hospital* OR ((health OR treatment OR care) NEAR/2 (center* OR centre*)))) |
| S5 | [S2] AND [S3] AND [S4] AND [S5] |
| S6 | ([S2] AND [S3] AND [S4] AND [S5]) AND yr(2010-2024) |

